# The effect of social restrictions, loss of social support, and loss of maternal autonomy on postpartum depression in 1 to 12-months postpartum women during the COVID-19 pandemic

**DOI:** 10.1101/2021.10.21.21265354

**Authors:** Kanami Tsuno, Sumiyo Okawa, Midori Matsushima, Daisuke Nishi, Yuki Arakawa, Takahiro Tabuchi

## Abstract

**Background:** This study focuses on postpartum women, who are one of the most vulnerable populations during the COVID-19 pandemic, aiming to reveal mental health consequences of social restrictions, loss of social support, or loss of autonomy.

**Methods:** A cross-sectional study for postpartum women was conducted in October 2020 (N = 600). The Edinburgh Postpartum Depression Scale (EPDS) was used to measure postpartum depression. The prevalence ratios were estimated by log-binomial regression models, adjusting for age, education, household income, residence area, parity, the timing of delivery, and a prior history of depression.

**Results:** The prevalence of postpartum depression was 28.7% (EPDS ≥ 9), 18.6% (≥ 11), and 13.1% (≥ 13). Social restrictions including cancellation of home visits by health care professionals, or cancellation of infant checkups or vaccinations, loss of support during pregnancy or after delivery including loss of opportunities to consult with health care professionals or friends, or cancellation of parents or other family members’ visits to support, and loss of autonomy about delivery or breastfeeding, were associated with postnatal depression.

**Conclusion:** About 30% of women who delivered and raised a baby during the COVID-19 pandemic had postpartum depression, which is much higher than a pre-pandemic meta-analysis. COVID-19 related social restrictions or loss of social support from healthcare professionals, family, and friends were significantly associated with postpartum depression. Also, loss of maternal autonomy in delivery and breastfeeding is associated with postpartum depression. The results indicate that both formal and informal support should not be limited to prevent postpartum depression during the pandemic.

## 1. Introduction

The coronavirus disease 2019 (COVID-19) pandemic has changed people’s daily lives at a global level with imposed social restriction on the citizens to contain an infection in addition to the spread of the virus itself. In this pandemic, one of the most vulnerable populations is postnatal women since most supporting systems or services have been canceled at birth facilities or public health centers under infection control. According to the survey of the Japanese Society of Obstetrics and Gynecology (JSOG) conducted in September 2020 (Komatsu et al., 2020), 92% of the facilities canceled birth and parenting classes, 80% prohibited birth attendance by partner, and 86% prohibited family, relatives, and friends visits during the postpartum hospital stay. Although such measures may contribute to lower risk of infection among perinatal women, it also exposes them to isolation and higher risk of postnatal depression (Viaux et al., 2020). Simultaneously, pregnant women felt a heavy burden not to be infected with SARS□CoV□2 because 66% and 80% of the facilities in Japan transferred pregnant women if they had an asymptomatic and symptomatic infection, respectively (Komatsu et al., 2020).

Generally, 10-20% of mothers have experienced postpartum depression. The recent meta-analysis has reported the global pooled prevalence of postpartum depression as 17.7% (Hahn-Holbrook et al., 2017). A systematic review and meta-analysis have reported the COVID-19 pandemic significantly increases the risk of anxiety among women during pregnancy and perinatal period (Hessami et al., 2020). So far, the percentages of pregnant women who obtained scores higher than 13 on the Edinburgh Postpartum Depression Scale (EPDS) were reported to be 17% in Japan (Matsushima and Horiguchi, 2020); 29.6% in China (Wu et al., 2020); 35.4% in Turkey (Durankus and Aksu, 2020); 37% in Canada (Lebel et al., 2020). However, the prevalence of postpartum depression among postpartum women who delivered during the COVID-19 pandemic has not been fully investigated and most of them are single-site studies (An et al., 2021; Ostacoli et al., 2020; Suzuki, 2020; Zanardo et al., 2020).

Lack of social support is one of the risk factors of postpartum depression (Beck, 2001). Social support is described as “the process by which interpersonal relationships promote and protect an individual’s well-being, particularly when that person is faced with stressful life circumstances” (Wills and Ainette, 2012). The COVID-19 pandemic is one of the stressful life circumstances for perinatal women. Moreover, under this situation, every kind of social support that they were supposed to obtain would have decreased due to the cancelation of parenting classes or hometown delivery. The recent study during the COVID-19 pandemic has reported that cancelation of planned informal support and not receiving informal childcare support are associated with depression among pregnant women (Matsushima and Horiguchi, 2020). Nonetheless, the effects of social restriction and a decrease of social support on postpartum depression have not been fully investigated.

In Japan, it is common for pregnant women to give birth in their hometown and stay at their parents’ house before and after delivery to receive support from them. This is so-called ‘Satogaeri bunben’ (‘Satogaeri’ means returning to the parents’ house and ‘bunben’ means delivery, hereafter referred to as “hometown delivery”), which is a traditional supporting system for perinatal women in Japan (Yoshida et al., 2001). However, pregnant or postpartum women had to cancel or refrain from such travel during the pandemic. For instance, the JSOG announced on their website that all pregnant women should refrain from having hometown delivery to prevent infection of SARS□CoV□2, as of April 21 (Japanese Society of Obstetrics and Gynecology, 2020). As a result, 17% of the birth facilities rejected pregnant women moving from other prefectures, according to the abovementioned JSOG survey (Komatsu et al., 2020). Even if a postpartum woman lives with her husband/partner, social support from him might not be enough because the time Japanese working fathers have engaged in household chores and childcare is only one-fifth of those of mothers, according to the 2020 Annual Report on the State of the Formation of a Gender Equal Society (Cabinet Office, 2020). Thus, the cancelation of hometown delivery exposes mothers to loss of social support and a heavy burden of household chores and child care, which may cause an adverse mental health condition among mothers.

The aim of the current study was therefore to investigate the effect of various social restrictions or loss of support on postpartum depression among women who delivered and raised a baby during the pandemic.

## 2. Methods

### Study design and participants

This is a part of the “Japan COVID-19 and Society Internet Survey (JACSIS),” an ongoing longitudinal, population-based internet-based questionnaire survey (Ikeda et al., 2021; Yamada et al., 2021). The study design details have been described elsewhere (Miyawaki et al., 2021). The JACSIS study consisted of 3 surveys with the following targets: general population (N = 28,000), pregnant and postpartum women (N = 1,000), and single mothers/fathers (N = 1,000). The study sample for each survey was retrieved from the pooled panels of an internet research agency (Rakuten Insight, Inc.), which had about 2.2 million panelists in 2019.

In this study, we used the baseline cross-sectional data of the pregnant and postpartum women survey which was collected on October 15-25, 2020. Four thousand three hundred seventy-three women who had given birth later than October 2019 or were expected to give birth by March 2021 were recruited from 21,896 eligible samples using simple random sampling. Data were collected until the time the target sample size reaches 1000 (participation rate, 22.9%) which comprised of the following 4 groups: women who delivered between October 2019 and March 2020 (n = 200), between April and May 2020 (n = 200), who have delivered later than June (n = 200), and who were pregnant at the time of the survey as well as expected to deliver by March 31, 2021 (n = 400). Among 1,000 participants, we analyzed the data of postpartum women who delivered a baby between October 2019 and October 2020 (n = 600).

### Measurements

#### Social restriction, loss of support, and loss of autonomy during pregnancy

We asked the participants if they experienced any kind of social restrictions (4 items), loss of social support (2 items), or loss of autonomy (1 item) during pregnancy (see Table 2). The response options were “yes” or “no.”

#### Social restrictions and loss of support after the delivery

We asked the participants if they experienced any kind of social restrictions or loss of social support after delivery using 6 and 5 items, respectively (see Table 3). The response options were “yes” or “no”.

### Postpartum depression

The Japanese version of the EPDS was used to measure postpartum depression (Cox et al., 1987; Okano, 1996). The EPDS consists of 10 items rated on a four-point Likert scale ranging from 0 to 3, with a total score between 0 and 30. The Japanese version of the EPDS has been reported good reliability and validity. In this study, the Cronbach alpha of EPDS was 0.86. Individuals who score 9 or more are considered at high risk of postpartum depression in Japan (Okano, 1996). In addition, we also investigated another definition of postpartum depression using EPDS, which is 11 or higher and 13 or higher, since a recent systematic review and meta-analysis reported a cutoff value of 11 or higher maximized combined sensitivity and specificity, and a cut-off value of 13 or higher was less sensitive but more specific (Levis et al., 2020).

### The timing of delivery

The timing of delivery was categorized into three: before the first state of emergency (October 2019 to March 2020), during the first state of emergency (April 2020 to May 2020), and after the first state of emergency (June 2020 to October 2020). On April 7, the Japanese government declared the first state of emergency in 7 prefectures including Tokyo. It has urged citizens to stay at home and to avoid unnecessary travel, particularly from cities to rural areas where many elderly people live (Looi, 2020). Following a further increase in untraceable cases, the declaration was expanded across the nation on April 16, with 13 major prefectures categorized as “prefectures under special precautions.” As the number of new cases began to fall, the declaration was lifted on May 25 in all regions. To investigate the effect of the state of emergency, we compared the number of mothers who experienced social restrictions and loss of social support by the timing of delivery.

### Confounders

Based on the reported predictors of postpartum depression (Beck, 2001) and the specific situation of COVID-19 measures, we measured potential confounders as follows: age (under 29, 30-34, 35-39, or over 40), residence area (prefectures under special precautions or others), education (high school or below, junior college/vocational school, or university or above), annual household income during the previous year (199 or less, 200-399, 400-599, 600-799, 800-999, 1,000 or more, or unknown [ten thousand Japanese yen]), having a partner/husband (yes or no), living with a partner/husband (yes or no), living with parents/parents in law (yes or no), parity (primipara, multipara), unplanned/unwanted pregnancy, and having depression (never, past, or current).

### Statistical analyses

Inaccurate responses include the following 3 definitions; those who selected “yes” to all disease lists (do you have the following diseases? (16 items listed below), those who selected “using some times or almost every day” to all items on alcohol product or drug use (Do you currently use alcohol products or drugs? [9 items]), and those who selected other than “D” out of “A B C D E” in the question of “please select the second option from the bottom.” We excluded these respondents with discrepancies or artificial/unnatural responses (n = 42, remaining n = 558).

First, we compared the EPDS scores and the socio-demographic characteristics. Then we compared the percentages who experienced social restrictions or loss of support during pregnancy or after delivery by the timing of delivery. Lastly, the prevalence ratios of postpartum depression (defined as having an EPDS score of 9+, 11+, and 13+) among those who had experienced social restrictions or loss of support during pregnancy or after delivery were estimated by a log-binomial regression model. The reason why this model was selected is in this study is that prevalence of postpartum depression was over 10% and the odds ratio could overestimate the prevalence ratio (Barros and Hirakata, 2003). In the series of analyses, we first adjusted age, education, household income, residence area, parity, and the timing of delivery. Subsequently, a prior history of depression was adjusted. The level of significance was 0.05 (two-tailed). The statistical analyses were conducted using SPSS 27.0 for Windows (IBM, Japan).

## 3. Results

### Characteristics of the participants

We analyzed 558 women after excluding 42 participants who provided inaccurate responses we mentioned above. Most participants were an age of 30-34, lived in prefectures under special precautions, graduated university, received 4.0-5.9 million yen as an annual household income, had a partner/husband, lived with a partner/husband, did not live with their parents or parents-in-law, were primipara, and never had depression previously (Table 1).

**Table 1.** Characteristics of the participants (N=558)

|  |  |  |  | Postnatal Depression (EPDS) |  |  |
| --- | --- | --- | --- | --- | --- | --- |
|  |  |  |  | ≥9 | ≥11 | ≥13 |
|  | n | % | Mean (SD) | n (%) | n (%) | n (%) |
| Age |  |  | 32.2 (4.1) | *p* = .428 | *p* = .129 | *p* = .114 |
| Under 29 | 140 | 25.1 |  | 47 (33.6) | 34 (24.3) | 26 (18.6) |
| 30-34 | 258 | 46.2 |  | 73 (28.3) | 43 (16.7) | 27 (10.5) |
| 35-39 | 134 | 24.0 |  | 34 (25.4) | 25 (18.7) | 18 (13.4) |
| Over 40 | 26 | 4.7 |  | 6 (23.1) | 2 (7.7) | 2 (7.7) |
| Residence area |  |  |  | *p* = .163 | *p* = .048 | *p* = .229 |
| Prefectures under special precautions | 376 | 67.4 |  | 115 (30.6) | 79 (21.0) | 54 (14.4) |
| Others | 182 | 32.6 |  | 45 (24.7) | 25 (13.7) | 19 (10.4) |
| Education |  |  |  | *p* = .031 | *p* = .069 | *p* = .327 |
| High school or below | 113 | 20.3 |  | 42 (37.2) | 28 (24.8) | 18 (15.9) |
| Junior college/vocational school | 160 | 28.7 |  | 36 (22.5) | 22 (13.8) | 16 (10.0) |
| University or above | 285 | 51.1 |  | 82 (28.8) | 54 (18.9) | 39 (13.7) |
| Annual household income during the previous year (Ten thousand Japanese yen) |  |  |  | *p* = .112 | *p* = .691 | *p* = .463 |
| 199 or less | 16 | 2.9 |  | 4 (25.0) | 2 (12.5) | 2 (12.5) |
| 200-399 | 74 | 13.3 |  | 30 (40.5) | 18 (24.3) | 14 (18.9) |
| 400-599 | 156 | 28.0 |  | 47 (30.1) | 28 (17.9) | 22 (14.1) |
| 600-799 | 120 | 21.5 |  | 28 (23.3) | 21 (17.5) | 15 (12.5) |
| 800-999 | 79 | 14.2 |  | 18 (22.8) | 12 (15.2) | 5 (6.3) |
| 1,000 or more | 49 | 8.8 |  | 11 (22.4) | 8 (16.3) | 6 (12.2) |
| Unknown | 64 | 11.5 |  | 22 (34.4) | 15 (23.4) | 9 (14.1) |
| Having a partner/husband |  |  |  | *p* = .325 | *p* = 1.000 | *p* = 1.000 |
| Yes | 554 | 99.3 |  | 158 (28.5) | 104 (18.8) | 73 (13.2) |
| No | 4 | 0.7 |  | 2 (50.0) | 0 (0.0) | 0 (0.0) |
| Living with a partner/husband |  |  |  | *p* = 1.000 | *p* = .485 | *p* = 1.000 |
| Yes | 544 | 97.5 |  | 156 (28.7) | 103 (18.9) | 72 (13.2) |
| No | 14 | 2.5 |  | 4 (28.6) | 1 (7.1) | 1 (7.1) |
| Living with parents/parents in law |  |  |  | *p* = .693 | *p* = 1.000 | *p* = 1.000 |
| Yes | 32 | 5.7 |  | 10 (31.3) | 6 (18.8) | 4 (12.5) |
| No | 526 | 94.3 |  | 150 (28.5) | 98 (18.6) | 69 (13.1) |
| Parity |  |  |  | *p* = .004 | *p* = .050 | *p* = .033 |
| Primipara | 294 | 52.7 |  | 60 (22.7) | 40 (15.2) | 26 (9.8) |
| Multipara | 264 | 47.3 |  | 100 (34.0) | 64 (21.8) | 47 (16.0) |

Table1 (cont.). Characteristics of the participants (N=558)
|  | n | % | Mean<br>(SD) | Postnatal Depression (EPDS) |  |  |
| --- | --- | --- | --- | --- | --- | --- |
|  |  |  |  | ≥9 | ≥11 | ≥13 |
|  | n | % |  | n (%) | n (%) | n (%) |
| Oct 2019 to Mar 2020 | 186 | 33.3 |  | 61 (32.8) | 42 (22.6) | 31 (16.7) |
| Apr 2020 to May 2020 | 188 | 33.7 |  | 46 (24.5) | 31 (16.5) | 19 (10.1) |
| Jun 2020 to Oct 2020 | 184 | 33.0 |  | 53 (28.8) | 31 (16.8) | 23 (12.5) |
| Unplanned/unwanted pregnancy |  |  |  | *p* = .895 | *p* = .878 | *p* = .600 |
| Yes | 82 | 14.7 |  | 136 (28.6) | 88 (18.5) | 61 (12.8) |
| No | 476 | 85.3 |  | 24 (29.3) | 16 (19.5) | 12 (14.6) |
| Having depression |  |  |  | *p* = .020 | *p* = .001 | *p* = .009 |
| Never | 505 | 90.5 |  | 137 (27.1) | 87 (17.2) | 60 (11.9) |
| Past | 43 | 7.7 |  | 17 (39.5) | 11 (25.6) | 9 (20.9) |
| Current | 10 | 1.8 |  | 6 (60.0) | 6 (60.0) | 4 (40.0) |
| Postnatal Depression (EPDS) |  |  |  |  |  |  |
| Total score (0-27) |  |  | 6.5 (5.2) |  |  |  |
| EPDS ≥9 |  |  |  |  |  |  |
| Yes | 160 | 28.7 |  |  |  |  |
| No | 398 | 71.3 |  |  |  |  |
| EPDS ≥11 |  |  |  |  |  |  |
| Yes | 104 | 18.6 |  |  |  |  |
| No | 454 | 81.4 |  |  |  |  |
| EPDS ≥13 |  |  |  |  |  |  |
| Yes | 73 | 13.1 |  |  |  |  |
| No | 485 | 86.9 |  |  |  |  |

EPDS: Edinburgh Postnatal Depression Scale; SD: Standard Deviation.

### Prevalence of postpartum depression

The prevalence of postpartum depression was 28.7% (EPDS ≥ 9), 18.6% (EPDS ≥ 11), and 13.1% (EPDS ≥ 13). Participants who lived in prefectures under special precautions, whose educational attainment was high school or low, multipara, and had experienced depression currently or past were more likely to have postpartum depression.

### Social restrictions, loss of support, and loss of autonomy during pregnancy and the timing of delivery

During pregnancy, 63.1% of participants had experienced cancellation of birth and parenting classes, 25.1% felt that she had no choice but to follow the doctor’s instructions without telling her preferences about the delivery and breastfeeding, 17.6% were unable to talk to a friend or acquaintance about problems during pregnancy, 10.2% gave up hometown delivery, and 5% reduced the number of prenatal checkups (Table 2). Overall, 1.6% were transferred because the hospital stopped accepting births or experienced that the hospital/clinic where the hometown delivery had been scheduled refused it.

**Table 2.**
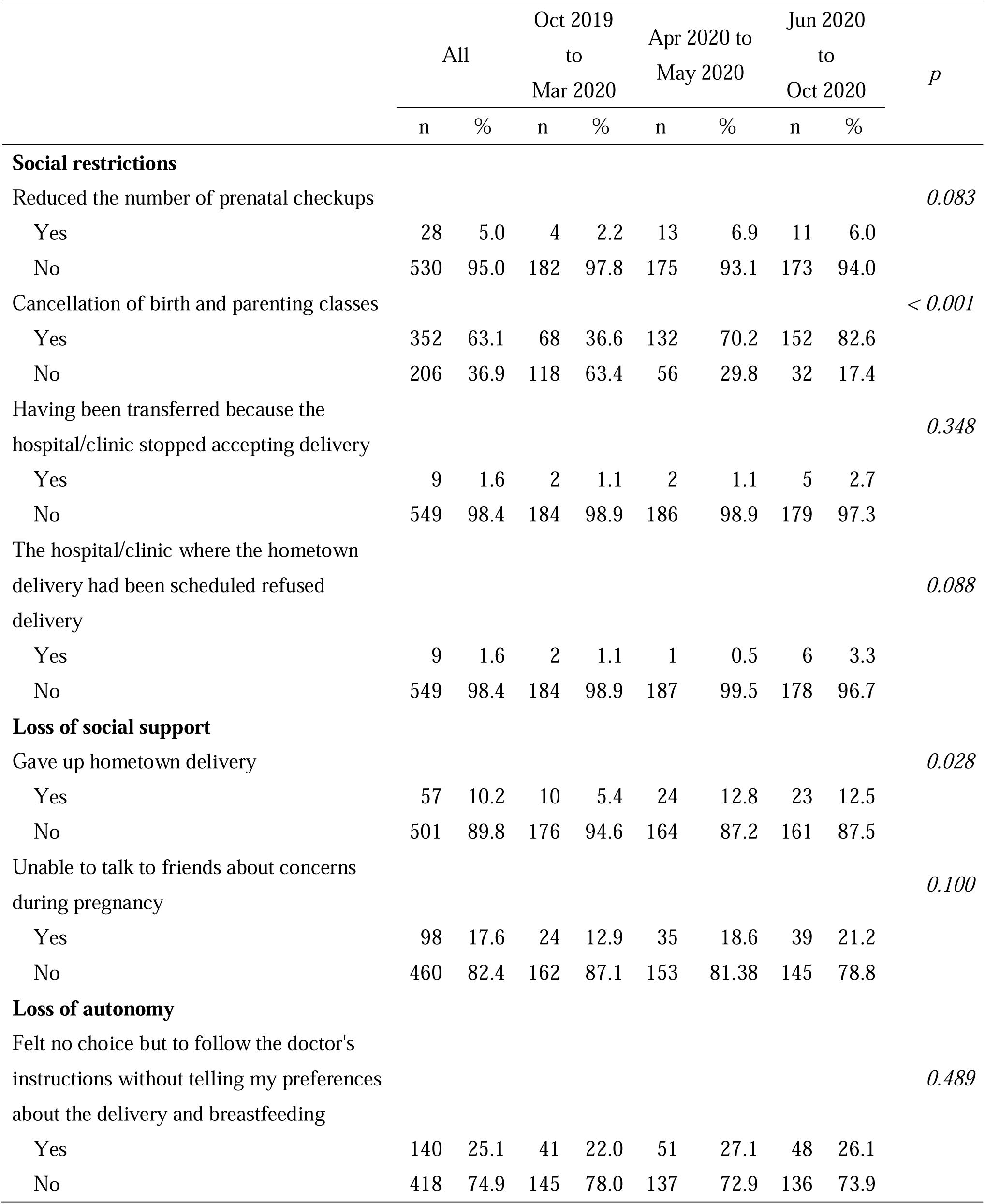
Social restrictions and loss of support during pregnancy and the timing of delivery

Compared to postpartum women who delivered a baby before the pandemic (October 2019 to March 2020), those who delivered during the pandemic (April 2020 to May 2020 [during the state of emergency] and June 2020 to October 2020) were more likely to unable to attend birth and parenting classes during pregnancy (70.2% and 82.6% VS 36.6%) or give up hometown delivery (12.8% and 12.5% VS 5.4%).

### Social restrictions or loss of support after delivery and the timing of delivery

After delivery, most postpartum women (73.1%) experienced the restriction of her family members’ visits during her hospital stay and the absence of her husband during childbirth (Table 3). In addition, 24.7% could not get her parents or other family members to come and help her after delivery, 22.2% felt that there were few opportunities for doctors, midwives, and nurses to teach her about childcare and breastfeeding, 14.3% were unable to talk to a friend or acquaintance about breastfeeding or childcare issues, 13.4% were unable to consult with a doctor or midwife about her health, and 13.3% voluntarily refrained from receiving home visits by public health nurses or midwives.

**Table 3.** Social restrictions and loss of support after childbirth and the timing of delivery

|  | All |  | Oct 2019 to<br>Mar 2020 |  | Apr 2020 to<br>May 2020 |  | Jun 2020 to<br>Oct 2020 |  | *p* |
| --- | --- | --- | --- | --- | --- | --- | --- | --- | --- |
|  | n | % | n | % | n | % | n | % |  |
| **Social restrictions** |  |  |  |  |  |  |  |  |  |
| No husband/partner attendance in the<br>delivery room despite my wish |  |  |  |  |  |  |  |  | 0.873 |
| Yes | 23 | 4.8 | 7 | 4.1 | 9 | 5.2 | 7 | 5.1 |  |
| No | 456 | 95.2 | 163 | 95.9 | 163 | 94.8 | 130 | 94.9 |  |
| Restrictions of family visits during the<br>hospital stay |  |  |  |  |  |  |  |  | < 0.001 |
| Yes | 408 | 73.1 | 51 | 27.4 | 181 | 96.3 | 176 | 95.7 |  |
| No | 150 | 26.9 | 135 | 72.6 | 7 | 3.7 | 8 | 4.3 |  |
| Not given home visits services by public<br>health nurses or midwives despite my<br>request |  |  |  |  |  |  |  |  | 0.887 |
| Yes | 24 | 4.3 | 7 | 3.8 | 9 | 4.8 | 8 | 4.3 |  |
| No | 534 | 95.7 | 179 | 96.2 | 179 | 95.2 | 176 | 95.7 |  |
| Voluntarily refrained from receiving<br>home visits by public health nurses or<br>midwives |  |  |  |  |  |  |  |  | < 0.001 |
| Yes | 74 | 13.3 | 21 | 11.3 | 39 | 20.7 | 14 | 7.6 |  |
| No | 484 | 86.7 | 165 | 88.7 | 149 | 79.3 | 170 | 92.4 |  |
| Not given infant health checkups and<br>vaccinations despite my request |  |  |  |  |  |  |  |  | 0.037 |
| Yes | 29 | 5.2 | 16 | 8.6 | 7 | 3.7 | 6 | 3.3 |  |
| No | 529 | 94.8 | 170 | 91.4 | 181 | 96.3 | 178 | 96.7 |  |
| Voluntarily refrained from having<br>infant checkups and vaccinations |  |  |  |  |  |  |  |  | 0.459 |
| Yes | 27 | 4.8 | 11 | 5.9 | 10 | 5.3 | 6 | 3.3 |  |
| No | 531 | 95.2 | 175 | 94.1 | 178 | 94.7 | 178 | 96.7 |  |

Table 3 (cont.). Social restriction and lack of support after childbirth and the timing of delivery
|  | All |  | Oct 2019 to<br>Mar 2020 |  | Apr 2020 to<br>May 2020 |  | Jun 2020 to<br>Oct 2020 |  | *p* |
| --- | --- | --- | --- | --- | --- | --- | --- | --- | --- |
|  | n | % | n | % | n | % | n | % |  |
| **Loss of social support** |  |  |  |  |  |  |  |  |  |
| Cancellation of parents or other family members' visits to support |  |  |  |  |  |  |  |  | 0.000 |
| Yes | 138 | 24.7 | 27 | 14.5 | 62 | 33.0 | 49 | 26.6 |  |
| No | 420 | 75.3 | 159 | 85.5 | 126 | 67.0 | 135 | 73.4 |  |
| Cancellation of planned stay at my parents'/parents in-laws' house |  |  |  |  |  |  |  |  | 0.003 |
| Yes | 60 | 10.8 | 9 | 4.8 | 29 | 15.4 | 22 | 12.0 |  |
| No | 498 | 89.2 | 177 | 95.2 | 159 | 84.6 | 162 | 88.0 |  |
| Few opportunities of being taught about childcare and breastfeeding by doctors, midwives, and nurses at hospital ward |  |  |  |  |  |  |  |  | < 0.001 |
| Yes | 124 | 22.2 | 20 | 10.8 | 63 | 33.5 | 41 | 22.3 |  |
| No | 434 | 77.8 | 166 | 89.2 | 125 | 66.5 | 143 | 77.7 |  |
| Unable to consult with doctors or midwives about my health despite my needs |  |  |  |  |  |  |  |  | 0.618 |
| Yes | 75 | 13.4 | 23 | 12.4 | 29 | 15.4 | 23 | 12.5 |  |
| No | 483 | 86.6 | 163 | 87.6 | 159 | 84.6 | 161 | 87.5 |  |
| Unable to consult with friends about breastfeeding or childcare issues despite my needs |  |  |  |  |  |  |  |  | 0.698 |
| Yes | 80 | 14.3 | 24 | 12.9 | 30 | 16.0 | 26 | 14.1 |  |
| No | 478 | 85.7 | 162 | 87.1 | 158 | 84.0 | 158 | 85.9 |  |

Compared to postpartum women who delivered a baby before the pandemic, those who delivered during the pandemic were more likely to experience the restriction of her family members’ visits during her hospital stay (96.3%/95.7% VS 27.4%), lack of support from health care professionals or her family members (33.5%/22.3% VS 10.8%), and cancellation of planned stay at her parents’ or parents-in-laws’ house (15.4%/12.0% VS 4.8%). Those who delivered their baby during the first state of emergency were more likely to voluntarily refrain from receiving home visits by public health nurses or midwives after delivery than those who delivered a baby before the pandemic or after the first state of emergency was lifted (20.7% VS 11.3%/7.6%). On the other hand, mothers who labored before the pandemic were more likely to experience cancellation of infant health checkups or vaccinations despite her request (8.6% VS 3.7%/3.3%).

### Social restrictions, loss of support, and loss of autonomy during pregnancy and postnatal depression

After adjusting for age, education, household income, residence area, parity, the timing of delivery, and a prior history of depression, loss of autonomy about delivery or breastfeeding and loss of opportunity to talk with friends about concerns during pregnancy were associated with postnatal depression (EPDS ≥ 11 and 13) (Table 4). Additionally, when an outcome was defined as EPDS ≥ 9, reducing the number of prenatal checkups and giving up hometown delivery were also significantly associated with postnatal depression.

**Table 4.**
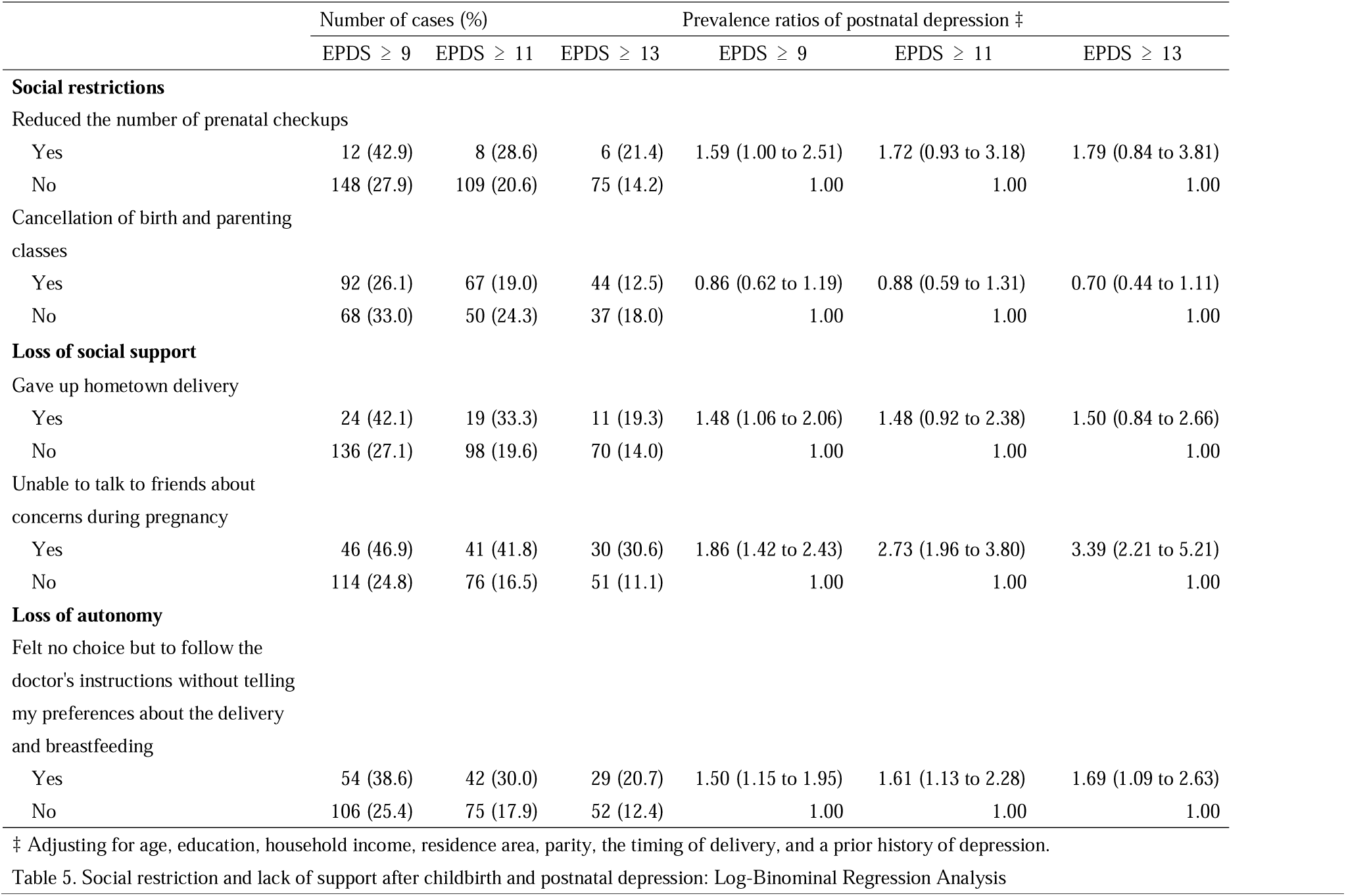
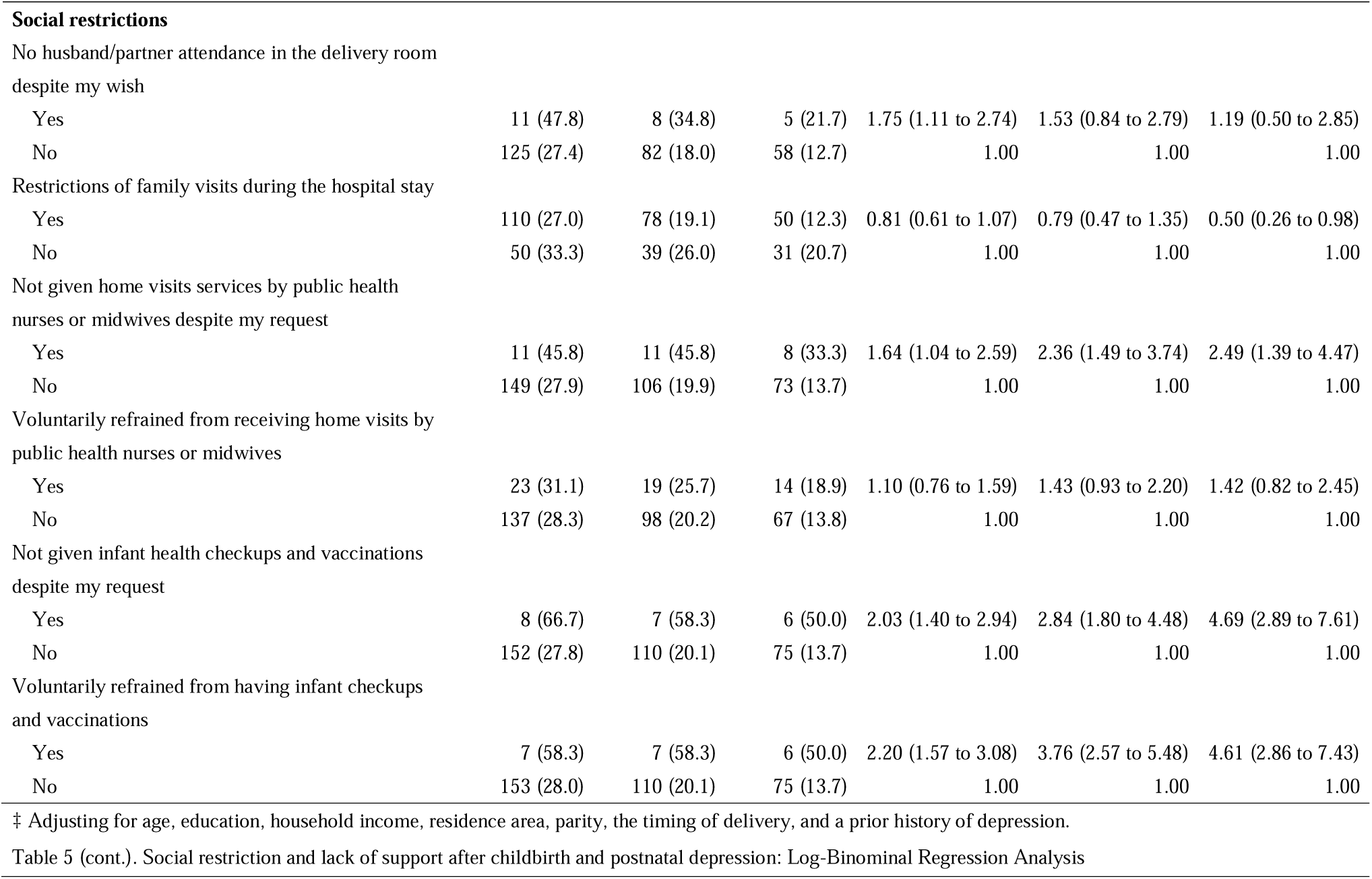
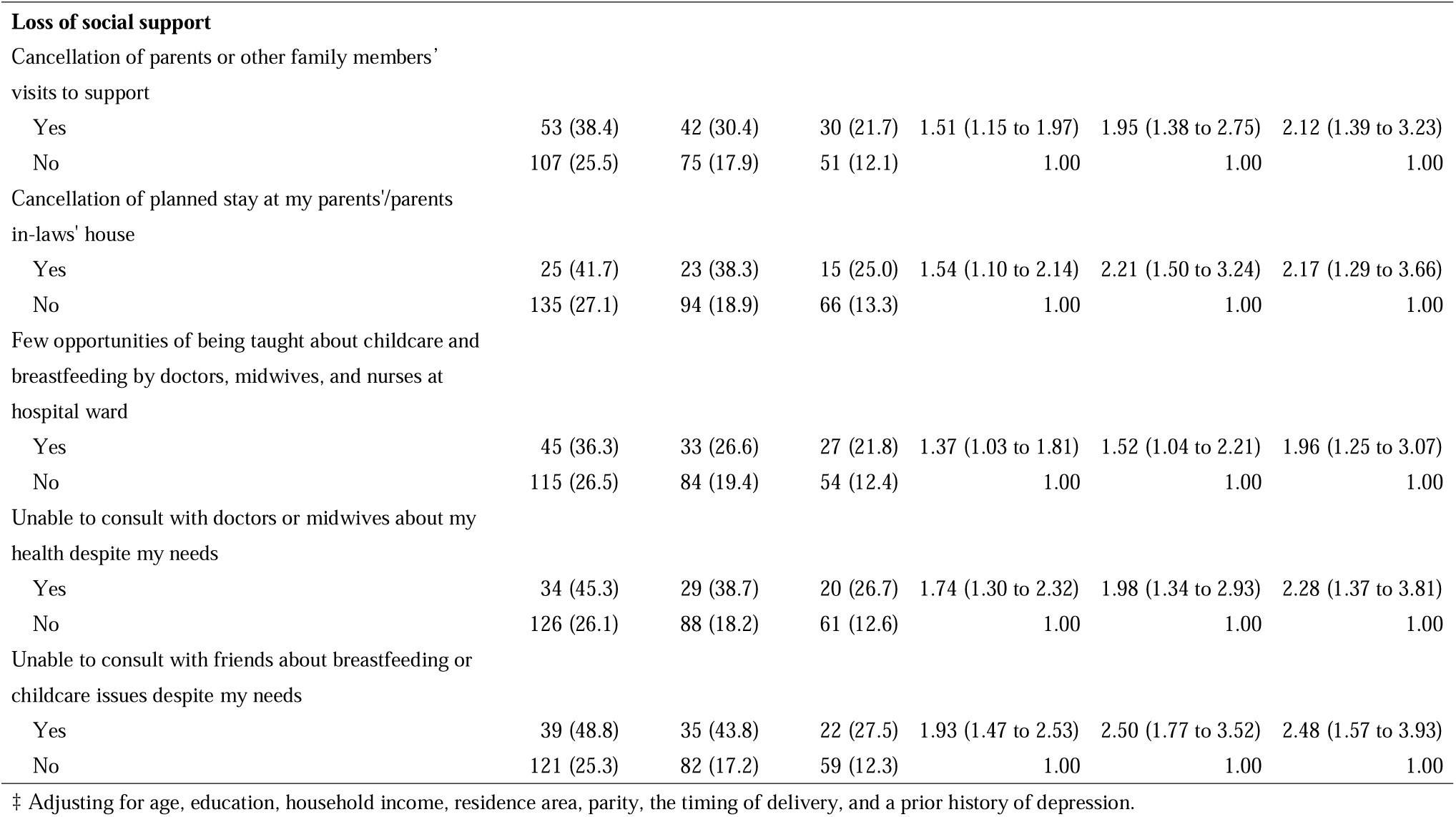
Social restrictions and lack of support during pregnancy and postnatal depression: Log-Binominal Regression Analysis

### Social restrictions or lack of support after delivery and postnatal depression

Among social restriction variables, loss of opportunities to receive home visits by health care professionals, loss of opportunities to receive infant checkups and vaccinations are significantly associated with all definitions of postnatal depression (EPDS ≥ 9, 11, and 13), after adjusting for age, education, household income, residence area, parity, the timing of delivery, and a prior history of depression (Table 5). The absence of a husband at childbirth was significantly associated with postnatal depression only in the EPDS ≥ 9 model. In contrast, restrictions of family visits during the hospital stay significantly associated with postnatal depression only in the EPDS ≥ 13 model.

All of the social support variables, loss of parents or family members’ support, cancelation of staying at her parents’ or parents-in-law’s house, few opportunities to learn about childcare and breastfeeding from health care professionals during the hospital stay, loss of opportunities to consult with health care professionals about own health, and loss of opportunities to talk with a friend/acquaintance about breastfeeding or childcare issues, are significantly associated with all definitions of postnatal depression.

## 4. Discussion

The current study investigated the effect of social restrictions or lack of social support on postpartum depression among women who delivered and raised a baby during the pandemic. Various social restrictions including loss of opportunities to receive infant health checkups and vaccinations, loss of support from healthcare professionals, family, or friends, and loss of autonomy regarding delivery or breastfeeding, were significantly associated with postnatal depression after adjusting for potential confounders. The study results indicate that both formal and informal support should not be limited to prevent postpartum depression during the COVID-19 pandemic.

Postpartum women who delivered a baby during the pandemic were more likely to experience social restrictions and loss of support during pregnancy and after delivery than those who delivered a baby before the pandemic. Under the birth facilities’ COVID-19 preventive measures (Japanese Society of Obstetrics and Gynecology, 2020), they lost chances of attending birth and parenting classes, consulting with health care professionals, or accepting family visits during the hospital stay, and had to cancel hometown delivery or a planned stay at her parents’/parents in-laws’ house. This is in line with the study that reported many pregnant women experienced these social restrictions during the pandemic (Matsushima and Horiguchi, 2020). These social restrictions would cause a loss of social support from health care professionals, family, and friends. For example, one-third of women answered they could not get support from their parents or other family members in this study. Although surveys of social restrictions were previously conducted for birth facilities (Komatsu et al., 2020) or pregnant women (Matsushima and Horiguchi, 2020), no study have asked postpartum women what kind of social restrictions, loss of support, or loss of autonomy they experienced during the pandemic. The results of this study expand the scientific evidence and indicate the COVID-19 preventive measures expose mothers to loss of social support.

Various COVID-19 related social restrictions and lack of social support were found to be associated with postnatal depression in our study. This is consistent with a previous study for pregnant women that reported the cancelation of planned informal support and not receiving informal childcare support associated with prenatal depression during the COVID-19 pandemic (Matsushima and Horiguchi, 2020), as well as the study reporting that a lack of social support is one of the risk factors of postpartum depression (Beck, 2001; Collins et al., 1993). In addition to this evidence, the current study revealed specific three types of variables during the pandemic associated with postpartum depression, 1) loss of support from health care professionals, 2) loss of support from family or friends, and 3) loss of maternal autonomy in delivery and breastfeeding.

Social support from health care professionals mainly consists of informational support (e.g., information that might be used to deal with stress) (Cohen et al., 1985) because they provide how to breathe during delivery or how to raise a child at prenatal checkups, hospital stay, infant checkups, etc. In this study, 20-30% of mothers who gave birth during the pandemic experienced few opportunities of being taught about childcare and breastfeeding by doctors, midwives, and nurses during the hospital stay and they were twice more likely to have postpartum depression than those who did not. This indicates loss of informational support could associate with postpartum depression. Although face-to-face communication is difficult during the pandemic, at least online consulting should be provided by healthcare professionals for postpartum women.

Support from family or friends can include emotional support (e.g., expression of caring), tangible or instrumental support (e.g., direct material aid including finance), or belonging support (e.g., having others to engage in social activities) (Cohen et al., 1985). Although we did not ask the respondents the details of their received support, mothers who experienced a loss of support from their parents or other family members or loss of opportunities to consult with friends about childcare were twice more likely to have postpartum depression. In other words, this indicates that informal support can prevent postnatal depression, in addition to formal support from health care professionals.

Around 25% of mothers experienced a loss of maternal autonomy in delivery or breastfeeding during the COVID-19 pandemic. As the longitudinal study has suggested (Gauthier et al., 2010), high levels of parental autonomous motivation (autonomy) predict lower levels of depressive symptoms among mothers. Autonomy is one of the basic psychological needs, according to Self-Determination Theory (SDT) (Deci and Ryan, 2000). SDT demonstrates a lack of needs satisfaction or the presence of need frustration may form a risk for adverse psychological health and several studies also suggest it determines maternal well-being or quality of parenting (Brenning and Soenens, 2017; Gauthier et al., 2010). Moreover, a study has reported a sense of control predicts depressive and anxious symptoms of mothers (Keeton et al., 2008). Thus our study result showing the association between a loss of autonomy in delivery or breastfeeding and postpartum depression is in line with those studies.

Contrary to our expectation, giving up hometown delivery was not significantly associated with postpartum depression when defined as EPDS ≥ 11 or 13, although the association was significant when using the definition of EPDS ≥ 9. This indicates the association between hometown delivery and postpartum depression is not strong. Although a non-significant association was against our expectation, this is in line with the cross-cultural study which reported hometown delivery itself did not lower the incidence of postnatal depression (Yoshida et al., 2001). When a woman delivered her baby in her hometown, she usually continues to stay at her parents’ home for several months. This helps her to receive supports from her parents but at the same time made her lose supports from her partner because they have to live separately. As Yoshida pointed out that this is a disadvantage of hometown delivery (Yoshida et al., 2001), which indicates hometown delivery may not always prevent postpartum depression.

While the cancellation of birth and parenting class did not associate with postpartum depression, restrictions of family visits during the hospital stay were negatively associated with postpartum depression. This might be due to the matching hypothesis of social support; social support buffers stress only when that support matches the stressful event or the receiver’s need (Cohen and Wills, 1985). Considering whether support responds to the individual’s needs or is perceived as supportive is more important than increasing the receipt of support (Wills and Shinar, 2000), it is case by case whether attending birth and parenting classes would reduce mothers’ stress and prevent postpartum depression. In other words, if birth and parenting classes match mothers’ needs, it may be beneficial for mothers’ mental health. On the contrary, mothers who experienced restrictions of family visits during the hospital stay were less likely to have postpartum depression. This might be because mothers were able to get rest during the hospital stay, thanks to no visitors. However, this association was only significant in EPDS ≥ 13 model so that our results should be generalized with caution.

When using the cutoff value of 9, the prevalence of postpartum depression was 28.7% in the current study, which is much higher than the prevalence of 14.4% among postpartum women during the pandemic (Suzuki, 2020) or a recent meta-analysis for Japanese women: 14.3% (Tokumitsu et al., 2020). On the other hand, when used higher cut off points (≥ 11 or 13), the prevalence of postpartum depression in the current study (18.6% or 13.1%) was much lower than other studies in the countries with high death tolls due to COVID-19: 28.6% or 44.2% (both EPDS ≥ 13) in Italy (Ostacoli et al., 2020; Zanardo et al., 2020); 56.9% (EPDS ≥ 10) in China (An et al., 2021). However, the prevalence was similar to other studies with the same definition (EPDS ≥ 13): 17% among Japanese pregnant women during the pandemic (Matsushima and Horiguchi, 2020), or 13-17% in meta-analyses before the pandemic (Hahn-Holbrook et al., 2017; O’hara and Swain, 1996). This indicates that mothers with a severe level of postpartum depression are not so many in Japan compared to other countries with high death tolls due to COVID-19 but those with a mild level of postpartum depression are prevalent, even where one of the smallest death toll was reported due to COVID-19 (8,929 as of 25 March 2021).

Surprisingly, the highest prevalence of postpartum depression in the current study was found among postpartum women who delivered a baby from October 2019 to March 2020, which is 7-12 months after birth. As reported by a recent meta-analysis (Tokumitsu et al., 2020), the prevalence of depression usually decreases in the postpartum period over time. For example, the period prevalence of postpartum depression (EPDS ≥ 9) was 15.1% within the first month, 11.6% in 1–3 months, 11.5% in 3–6 months, and 11.5% in 6–12 months after birth (Tokumitsu et al., 2020). Mothers who delivered a baby from October 2019 to March 2020 had to raise a baby during the state of emergency (April-May 2020). Under the state of emergency, even going outside for walking was hesitant because of high social pressure to stay home. The elementary schools or nursery schools were closed and their husbands started to work from home so that mothers had to take care of their babies and husbands, which increased mothers’ demands and raised mothers’ anxiety on how to raise their children (Takaku and Yokoyama, 2021). This can influence on the high prevalence of postpartum depression among 7-12 months postpartum women in the current study. The COVID-19 pandemic may have changed the pre-pandemic trend of postpartum depression so that healthcare professionals should provide long-term care for postpartum mothers.

## 5. Limitations

The current study has several limitations. First, we cannot determine the causal relationship due to a cross-sectional study design for some incidents. Although participants’ experiences during pregnancy or the hospital stay cannot be changed because this study limited postpartum women, the variables about infant checkups and vaccinations or social supports from health care professionals, family, and friends may be affected by psychological health status. We are planning a follow-up survey so that this will be solved shortly. Second, there might be a sampling bias due to the nature of an online survey. Also, since the participants had to answer many questions, some postpartum women with severe depression were unable to respond, which may underestimate the prevalence of postpartum depression. A longitudinal study with a large sample is needed to clarify the association between social restrictions and postpartum depression in the COVID-19 pandemic.

## 6. Conclusions

About 30% of women who delivered and raised a baby during the COVID-19 pandemic had postpartum depression, which is much higher than a pre-pandemic meta-analysis. COVID-19 related social restrictions or loss of social support from healthcare professionals, family, and friends significantly associated with postpartum depression. Also, loss of maternal autonomy in delivery and breastfeeding is associated with the risk of postpartum depression. The results indicate that both formal and informal support should be not be limited to prevent postpartum depression during the pandemic.

## Data Availability

The data used in this study are not available in a public repository because they contain personally identifiable or potentially sensitive patient information. Based on the regulations for ethical guidelines in Japan, the Research Ethics Committee of the Osaka International Cancer Institute has imposed restrictions on the dissemination of the data collected in this study. All data inquiries should be addressed to the person responsible for data management, Dr. Takahiro Tabuchi at the following e-mail address.

## Author Statement

### Contributors

Dr. Tsuno, Dr. Okawa, and Dr. Tabuchi designed the study, wrote the protocol, and collected the data. Dr. Tsuno undertook the statistical analysis and wrote the first draft of the manuscript. All authors significantly contributed to the interpretation of our data and revising the manuscript. All authors have approved the final manuscript.

## Acknowledge

We thank all researchers who participated and provided supports to the JACSIS study.

## Ethical issues

The study protocol was reviewed and approved by the Research Ethics Committee of the Osaka International Cancer Institute (approved on June 19, 2020; approval number 20084). All participants provided web-based informed consent before responding to the online questionnaire. A credit point known as “Epoints,” which could be used for internet shopping and cash conversion, was provided to the participants as an incentive.

